# Adverse Outcomes in Pulmonary Tuberculosis patients on Retreatment Regimen: Evaluation of Pharmacokinetic Estimates as Risk Indicators

**DOI:** 10.1101/2021.08.11.21261879

**Authors:** Anant Mohan, Anuj Bhatnagar, Tarang Gupta, Sunita Kanswal, Ujjalkumar Das, Thirumurthy Velpandian, Randeep Guleria, Urvashi B. Singh

## Abstract

Patients with pulmonary tuberculosis (PTB) who fail therapy or develop a relapse are initiated on re-treatment regimen. These patients are known to have adverse outcomes. This study aimed to determine the role of plasma levels of Anti-tubercular drugs in treatment outcome. Plasma levels of retreatment regimen drugs [Isoniazid(INH), Rifampicin(RIF), Pyrazinamide(PZA), Ethambutol(EMB), and Streptomycin(STM)] were compared between treatment responsive/cured and treatment failure/not-cured patients. Plasma drug levels were analysed by LC-MS/MS at different time points in 134 PTB patients on retreatment regimen. Of the 134 subjects, 108 were cured, 17 patients developed multi-drug resistant TB (MDR-TB), and 9 patients remained smear positive at treatment completion (8 months). The two-hour plasma levels (C_2hr_) (geometric mean) were lower in ‘Not Cured’ subjects compared to ‘Cured’ subjects Notably, in the 26 ‘Not Cured’ subjects, C_2hr_ plasma levels after first dose at Day0 were significantly low (INH: 0.86 vs 2.94 µg/ml p≤0.002, RIF: 0.56 vs 2.55 µg/ml p≤0.003, PZA: 1.85 vs 26.58 µg/ml p≤0.000 and EMB: 0.72 vs 1.53 µg/ml p≤0.010). In contrast, STM levels were higher (31.84 vs 18.08 µg/ml p ≤0.007). Based on ROC analysis of the data, therapeutic indicator values for successful treatment outcome were C_2hr_ plasma levels of 10.6 µg/ml for PZA, 1.14 µg/ml for RIF, 1.86 µg/ml for INH and 1.24 µg/ml for EMB. Therapeutic failure in PTB patients on retreatment regimen is associated with lower plasma drug levels. Therapeutic drug monitoring would prove useful to maintain drug levels above the minimum cut-off levels for obtaining favourable clinical outcome.

## Introduction

In cases of relapse after a successful anti-tubercular therapy (ATT), loss to follow-up or treatment failure, and in others who have undergone previous treatment to TB (1), the standardised retreatment regimen consisting of Isoniazid(INH/H), Rifampicin(RIF/R), Pyrazinamide (PZA/Z), Ethambutol(EMB/E) and Streptomycin(STM/S) for 2 months, followed by HRZE for 1 month and HRE for 5 months (2)is shown to render poor outcomes(3-5) and one of the reasons postulated for this retreatment failure in patients with TB is the sub-therapeutic ATT drug levels or their low bioavailability (6-10) that can lead to slower treatment response, acquired drug resistance, and relapse (11, 12).

Monitoring of plasma levels of drugs to tailor patient’s regimen, in a strategy called as ‘Therapeutic Drug Monitoring’ (TDM) has shown promising results in reaching requisite treatment goals in PTB patients (13, 14). Although TDM has been shown to predict clinical outcomes in treatment-naive PTB (12), no study has exclusively monitored drug levels in PTB on a retreatment regimen. TDM may have important implications for improving outcomes in this group of patients. To address this, we evaluated plasma levels of 5 ATT drugs on Day0 (0 and 2hrs after first dose) and after 2, 3, 5 & 8 months of initiation of treatment in subjects who were prescribed PTB retreatment regimen.

## Results

### Patients, Treatment Regimens and Outcomes

A total of 134 pulmonary TB patients with past history of TB and ATT medication were enrolled in the study. One hundred twenty one (90.3%) patients agreed to the intermittent ATT schedule. (DOTS as per RNTCP guidelines), while 13 (9.7%) preferred the daily regimen. The demographic details and clinical history of the patients is shown in Table 1 and microbiological testing data is shown in Table 2.

**Table 1:**
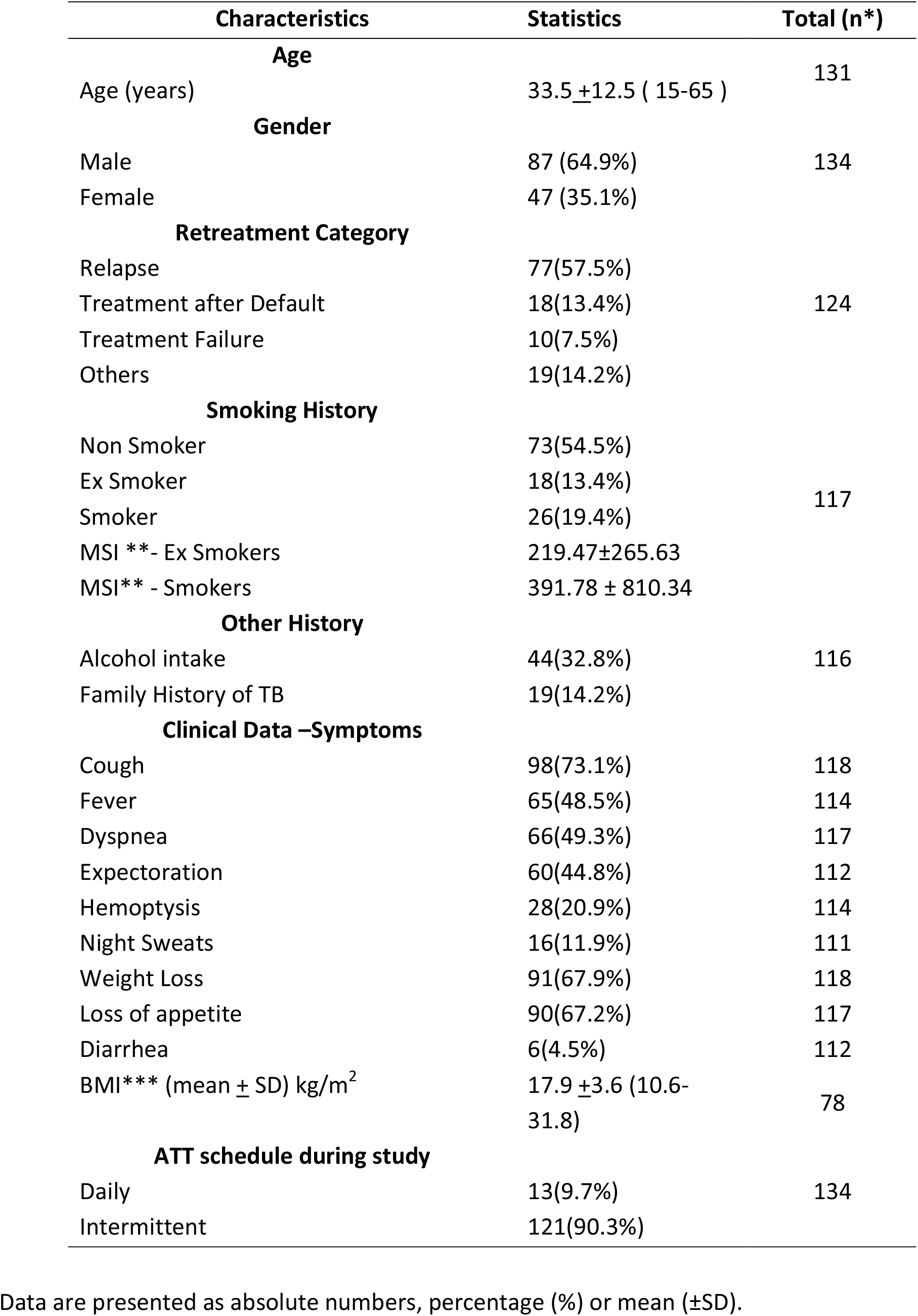
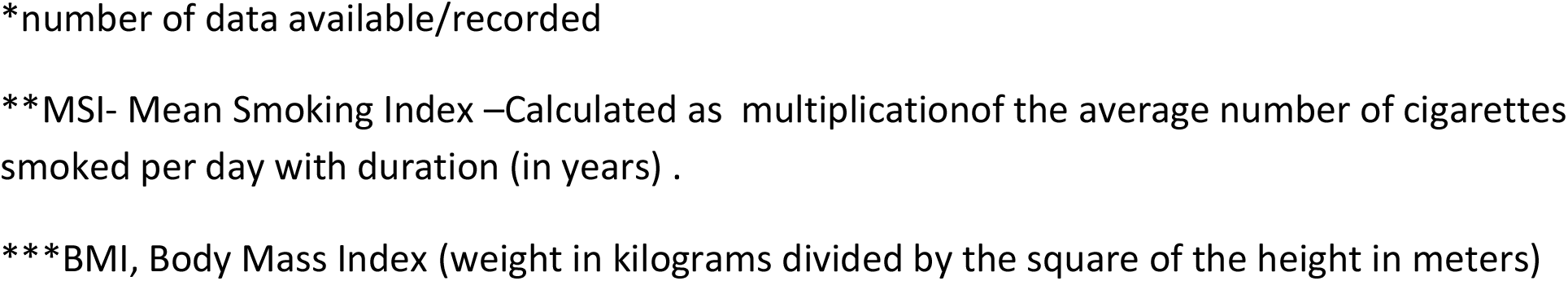
Basic Demographic and Clinical Details of Study Population.

| Characteristics | Statistics | Total (n*) |
| --- | --- | --- |
| Age |  |  |
| Age (years) | 33.5 ±12.5 ( 15-65 ) | 131 |
| Gender |  |  |
| Male | 87 (64.9%) | 134 |
| Female | 47 (35.1%) |  |
| Retreatment Category |  |  |
| Relapse | 77(57.5%) | 124 |
| Treatment after Default | 18(13.4%) |  |
| Treatment Failure | 10(7.5%) |  |
| Others | 19(14.2%) |  |
| Smoking History |  |  |
| Non Smoker | 73(54.5%) | 117 |
| Ex Smoker | 18(13.4%) |  |
| Smoker | 26(19.4%) |  |
| MSI **- Ex Smokers | 219.47±265.63 | |
| MSI** - Smokers | 391.78 ± 810.34 | |
| Other History |  |  |
| Alcohol intake | 44(32.8%) | 116 |
| Family History of TB | 19(14.2%) |  |
| Clinical Data –Symptoms |  |  |
| Cough | 98(73.1%) | 118 |
| Fever | 65(48.5%) | 114 |
| Dyspnea | 66(49.3%) | 117 |
| Expectoration | 60(44.8%) | 112 |
| Hemoptysis | 28(20.9%) | 114 |
| Night Sweats | 16(11.9%) | 111 |
| Weight Loss | 91(67.9%) | 118 |
| Loss of appetite | 90(67.2%) | 117 |
| Diarrhea | 6(4.5%) | 112 |
| BMI*** (mean ± SD) kg/m<sup>2</sup> | 17.9 ±3.6 (10.6-31.8) | 78 |
| ATT schedule during study |  |  |
| Daily | 13(9.7%) | 134 |
| Intermittent | 121(90.3%) |  |

Data are presented as absolute numbers, percentage (%) or mean ( ± SD).
\*number of data available/recorded
\*\*MSI- Mean Smoking Index –Calculated as multiplication of the average number of cigarettes smoked per day with duration (in years) .
\*\*\*BMI, Body Mass Index (weight in kilograms divided by the square of the height in meters)

**Table 2:**
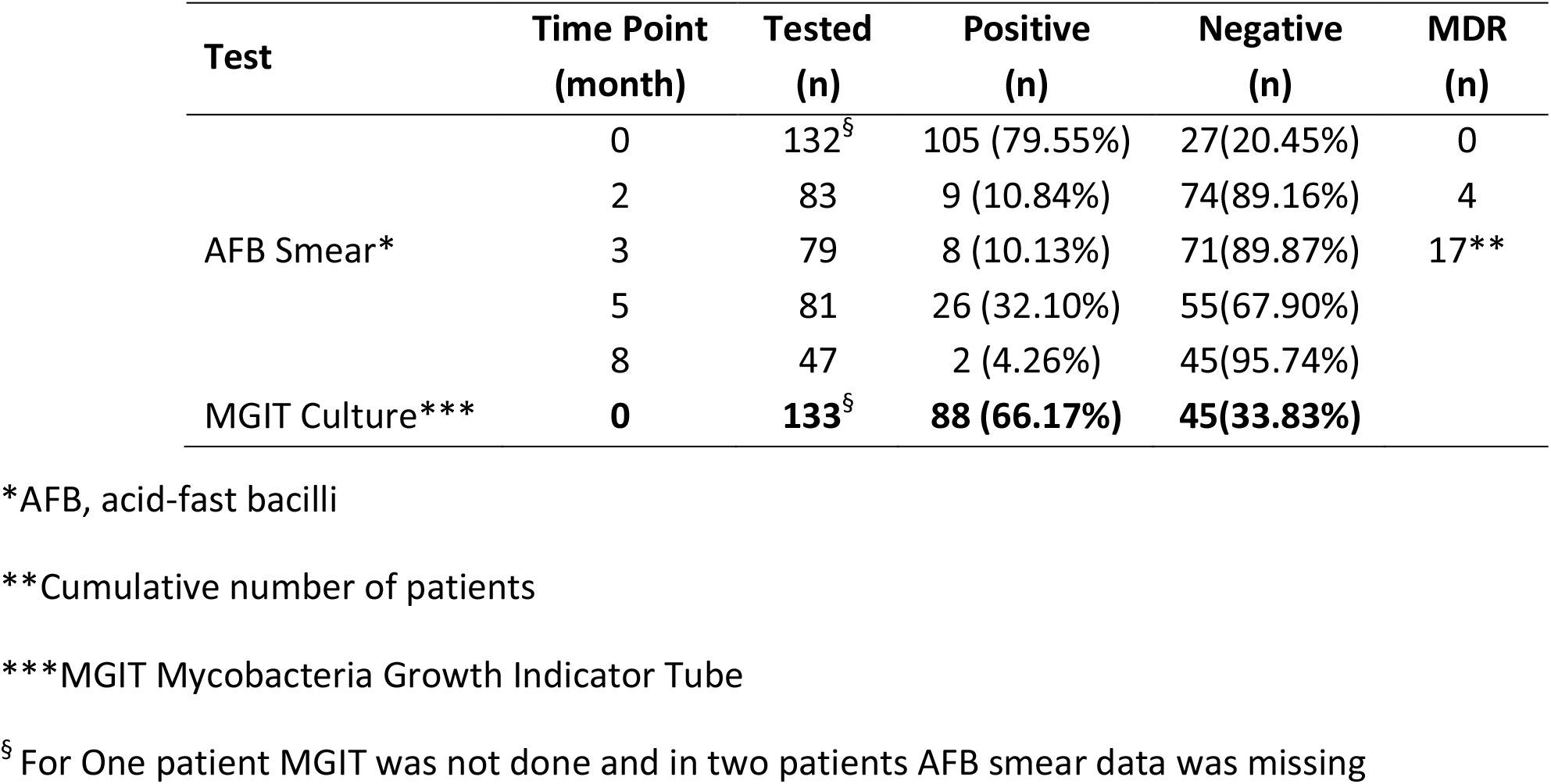
PTB DOTS Category II Patients – Microbiological Diagnosis Details.

| Test | Time Point<br>(month) | Tested<br>(n) | Positive<br>(n) | Negative<br>(n) | MDR<br>(n) |
| --- | --- | --- | --- | --- | --- |
| AFB Smear* | 0 | 132<sup>§</sup> | 105 (79.55%) | 27(20.45%) | 0 |
|  | 2 | 83 | 9 (10.84%) | 74(89.16%) | 4 |
|  | 3 | 79 | 8 (10.13%) | 71(89.87%) | 17** |
|  | 5 | 81 | 26 (32.10%) | 55(67.90%) |  |
|  | 8 | 47 | 2 (4.26%) | 45(95.74%) |  |
| MGIT Culture*** | 0 | 133<sup>§</sup> | 88 (66.17%) | 45(33.83%) |  |

\*AFB, acid-fast bacilli
\*\*Cumulative number of patients
\*\*\*MGIT Mycobacteria Growth Indicator Tube
<sup>§</sup> For One patient MGIT was not done and in two patients AFB smear data was missing

All 134 patients had a confirmed drug-susceptible PTB at the start of treatment. Of these, 108 (80.5%) patients were declared cured of TB and classified as ‘Cured’. Nine patients (6.72%) were not TB-free as per microbiological testing or physical examination. Seventeen patients (12.7 %) developed Rif resistance (RR) MDR (Multi-drug-resistant TB) by the end of third month of therapy and were referred for MDR regimen. Taken together, these 26 patients were categorized as ‘Not Cured’.

### Plasma Drug Levels

Plasma levels for H, R, Z, E and S were estimated in all patients for time points day0, followed by month 2, 3, 5 and 8 for H, R, E and day0, 2^nd^ month for Z and on day0 for S.

In the 17 patients who developed RR, plasma levels for H, R, Z, E and S were estimated until 2^nd^ and 3^rd^ month depending on time of MDR diagnosis.

A comparison of drug levels between ‘cured’ and ‘not cured’ patient groups according to timelines during the treatment is shown in table 3 and figure 1a. Statistically, plasma levels of isoniazid were significantly higher at 0, 2^nd^, and 5^th^ month time points, rifampicin levels were significantly higher at day0, comparable at month 2^nd^ and higher at 3^rd^, 5th and 8^th^ months.

**Table 3:**
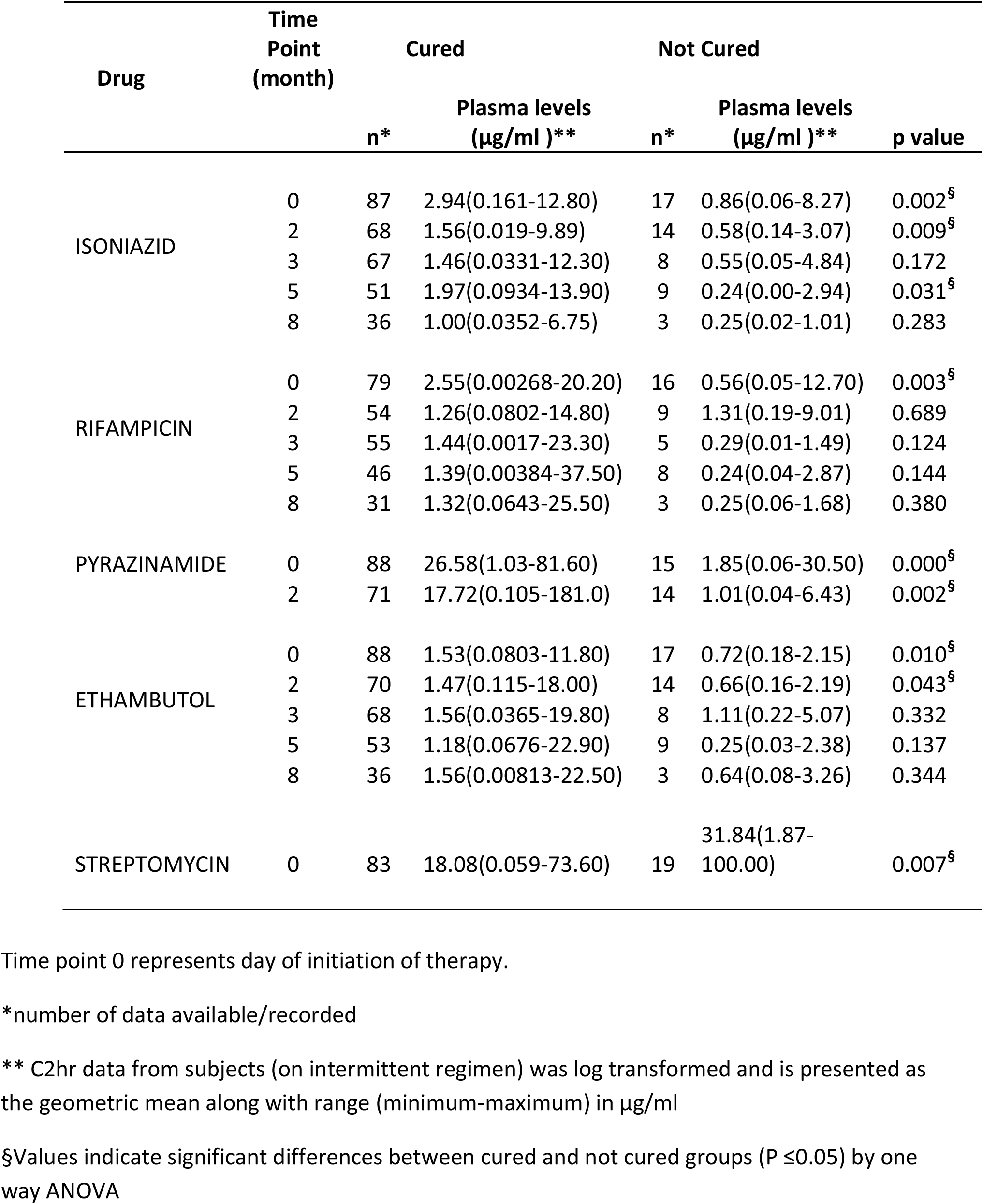
Plasma drug levels at 2 hrs post administration of ATT in DOTS category II PTB patients.

**Figure 1:**
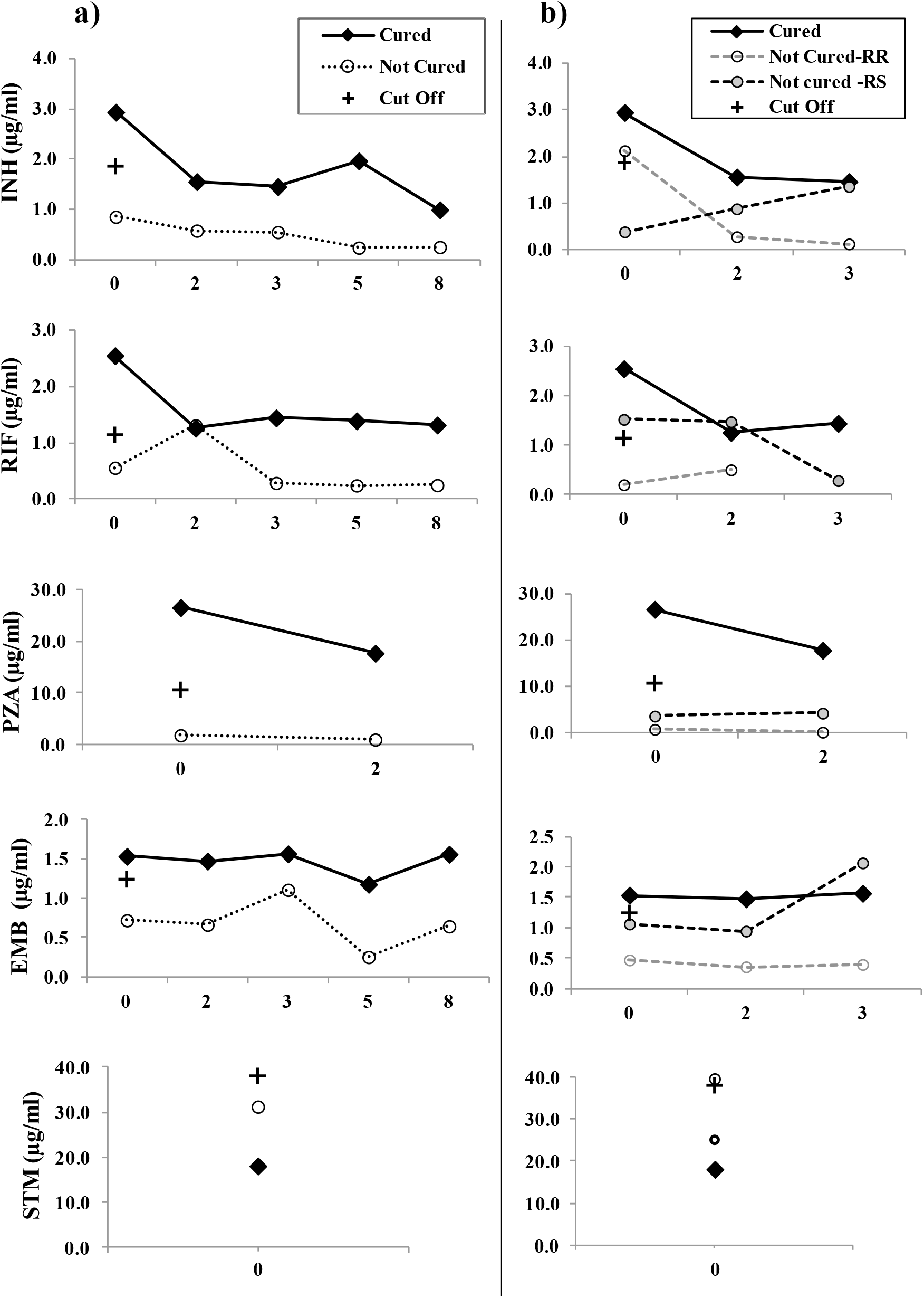
Serum levels (ug/ml) at different time points for Category II retreatment regimen drugs in PTB patients. a). ATT Plasma levels in Cured and Not Cured PTB patient groups. **b)** ATT Plasma levels in Cured, Rifampicin resistant (RR)MDR and Rifampicin sensitive (RS)Not Cured-groups. X axis shows time points of drug administration i.e. day 0, 2 month, 3 month, 5 month and 8 month (0, 2, 3, 5 and 8 -fig1a) or day 0,2^nd^ month and 3^rd^ month (0, 2 and 3 -fig1b). Y-axis shows geometric mean of plasma drug concentrations (µg/ml) of INH, RIF, PZA, EMB and STM as measured by HPLC-MS in patient blood samples after 2hrs of drug ingestion on day 0. Pyrazinamide is shown for 2 months and streptomycin C_2hr_ levels are shown for day 0 time point as per the treatment regimen. Solid line shows data from Cured subjects and dotted line shows data from Not-cured patients. **+** represents therapeutic indicator levels i.e. 0day-C_2hr_ levels required for a successful treatment outcome from cut off predicted from ROC analysis. Statistical significance is as shown in Table 3.

Pyrazinamide and ethambutol levels were significantly higher at both Day0 and 2^nd^ months. In contrast to the above four drugs, streptomycin levels were found to be significantly higher on day0 in the ‘Not Cured’ category of patients. Of note, all thirteen patients in the study who had been on daily dosing regimen, were cured after the therapy, while all patients who acquired drug resistance or failed treatment had been on intermittent therapy.

### Pharmokinetic variability in patient population

Plasma concentrations were found to be widely distributed in the patients. Overall, the highest drug concentrations were attained on day0-C_2hr_ time-point after which most peaks declined with time. Compared to the reference values for adults (15) (16), lower average (GM) drug plasma levels were prevalent at all time points in our study population (Table 3, figure 2). 22.3 % (27/121) of patients had plasma concentrations within the reference target range. Only 4/26 ‘not cured’ patients had drug levels within these target ranges and each of these patients achieved ‘within range’ plasma level for a single drug only.

**Figure 2.**
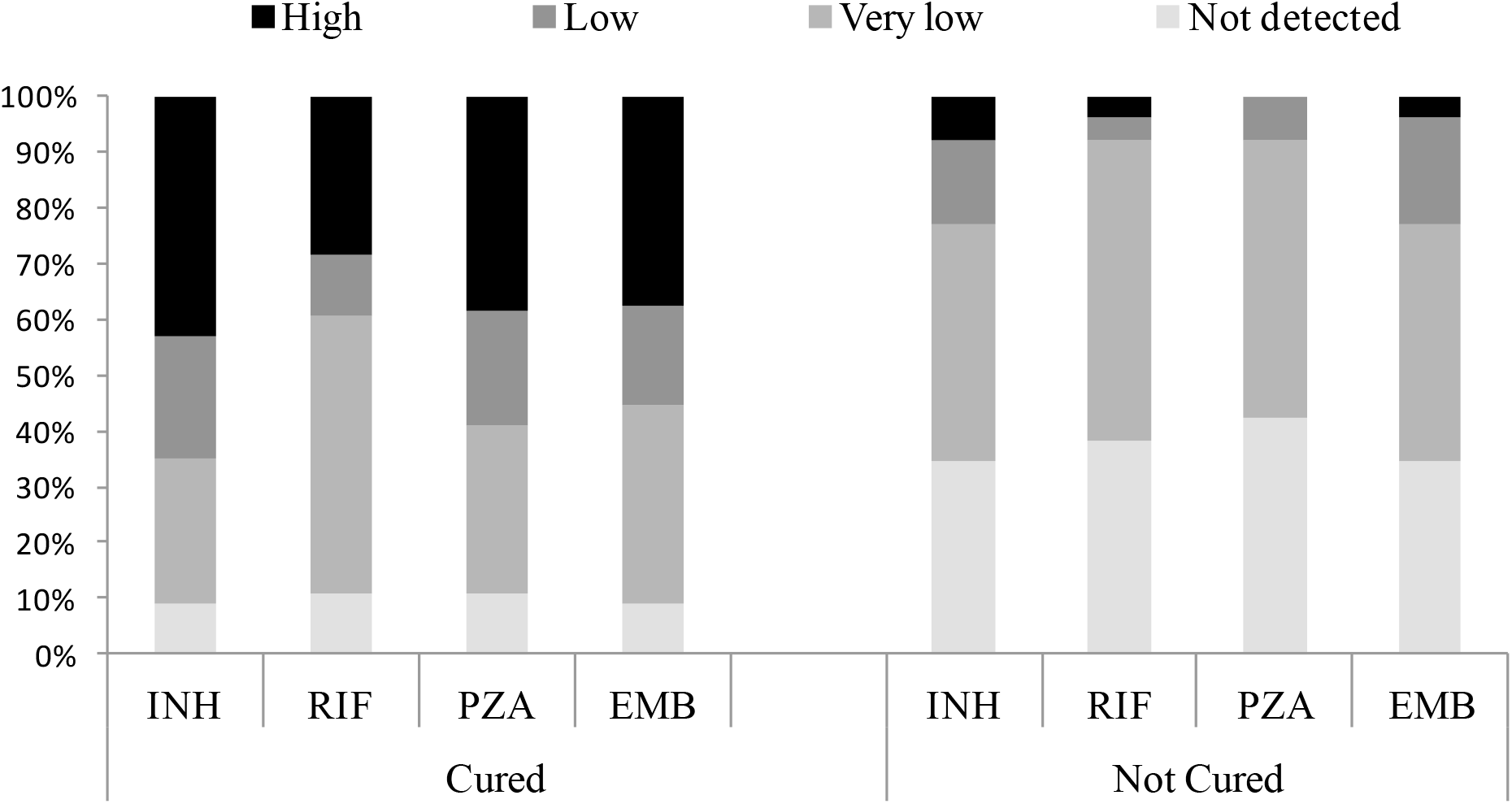
Pharmacokinetic variability of isoniazid, rifampicin, pyrazinamide and ethambutol in PTB patients on retreatment regimen at Day0-C_2hr_. Figure shows the percentage of patients with very low, low and target (high) serum concentrations at C2_hr_ on Day0 of drug dose administration in ‘cured’ and ‘not cured’ groups for isoniazid, rifampicin, pyrazinamide and ethambutol. Values are in comparison to adult reference ranges as reported previously (17) (18) Target ranges: H-3–6 µg/ml, R-8–24 µg/ml, Z-20-50 µg/ml and E-2–6 µg/ml (17) Low plasma concentrations: H < 3µg/ml, R < 8µg/ml, Z < 35 µg/ml and E < 2 µg/ml. Very low plasma concentrations: H < 1.5µg/ml, R < 4µg/ml, Z < 20 µg/ml and E < 1 µg/ml (18)

### Area under the ROC curve and predictor values for C2_hr_plasma ATT levels

Plasma drug levels were significantly lower in ‘Not Cured’ group when compared to the ‘Cured Group’ for all ATT drugs at C2_hr_ on day0. Hence we analysed the ROC curves for this time point (Table4, Figure3). The area under the ROC curve were the highest for PZA (0.924) followed by INH (0.788) and RIF (0.77). Optimal cut-off values that predict therapeutic indicator drug levels (minimum levels of drugs that when achieved in patient blood would likely predict a positive treatment outcome) are shown in Table 4. PZA (10.6 µg/ml, 86% sensitivity, 87% specificity) emerged as the best discriminating therapeutic indicator for treatment success in this group of patients (Figure 4).

**Table 4.** Receiver operator characteristics (ROC) values for plasma levels of ATT in PTB patients on Retreatment regimen

| ATT Drug | n | Area under the Curve | [ 95% CI ] | Optimal Cut off *<br>(µg/ml) | Sensitivity | Specificity |
| --- | --- | --- | --- | --- | --- | --- |
| ISONIAZID | 104 | 0.788 | [0.67-0.90] | 1.86 | 0.71 | 0.82 |
| RIFAMPICIN | 95 | 0.770 | [0.65-0.89] | 1.14 | 0.77 | 0.75 |
| PYRAZINAMIDE | 103 | 0.924 | [0.84-1.00] | 10.6 | 0.86 | 0.87 |
| ETHAMBUTOL | 105 | 0.712 | [0.60-0.83] | 1.24 | 0.60 | 0.76 |
| STREPTOMYCIN | 102 | 0.684 | [0.54-0.83] | 38.0 | 0.63 | 0.64 |

Table shows accuracy of area under the curve for C2hr plasma levels of ATT in PTB patients on retreatment regimen
\*Optimal cut off values (column 4) or Therapeutic indicator levels (minimum plasma drug concentrations that predict a positive treatment outcome ('Cured') in the study population).

**Figure 3:**
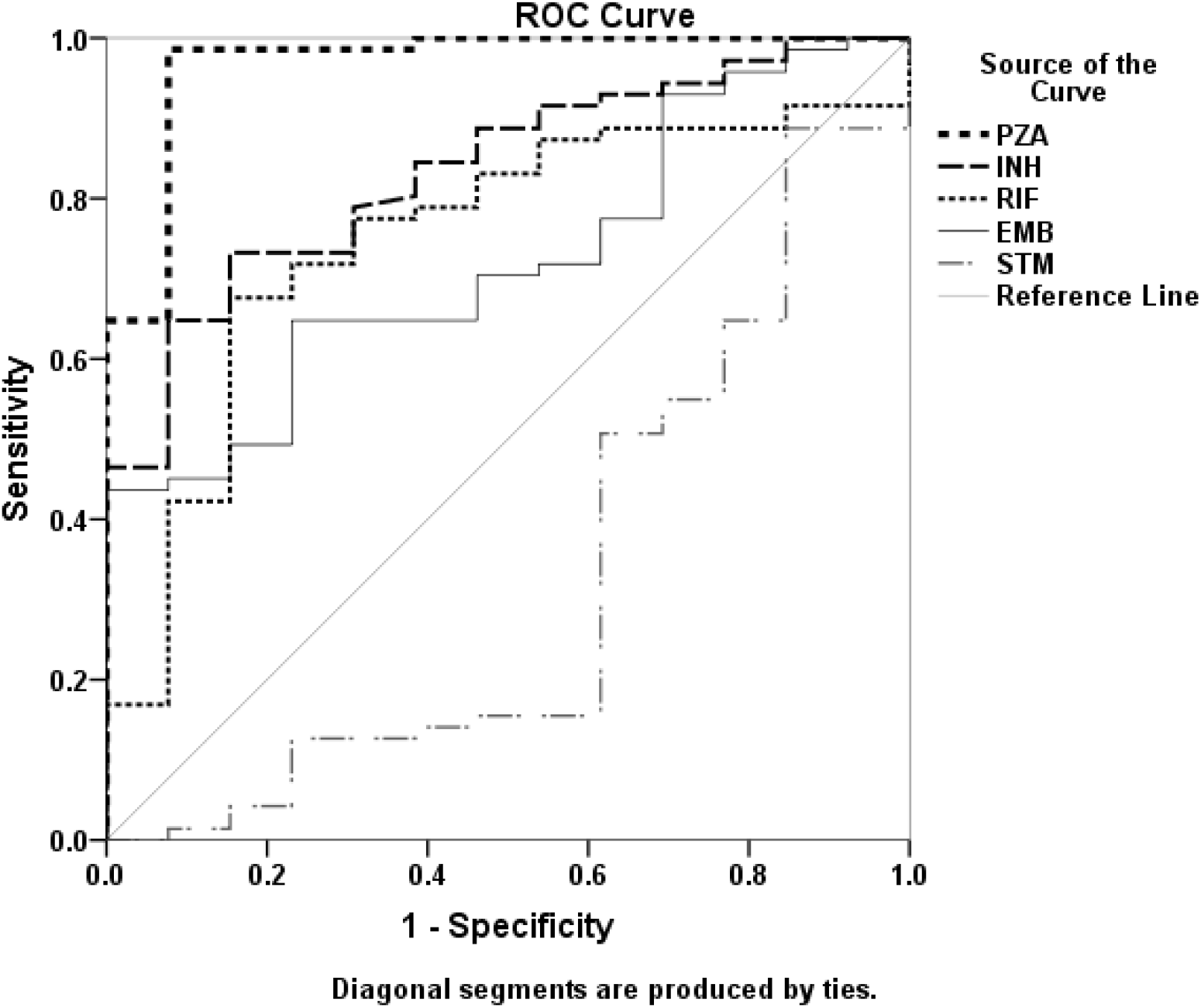
ROC Analysis of ATT drugs in PTB patients onretreatment regimen. Figure shows ROC curve analysis of Day0-C_2hr_ plasma levels of ATT-drugs, isoniazid (INH), rifampicin (RIF), pyrazimanide (PZA), ethambutol (EMB) and streptomycin (STM) in PTB patients on retreatment regimen. Values are as given in table 4.

**Figure 4:**
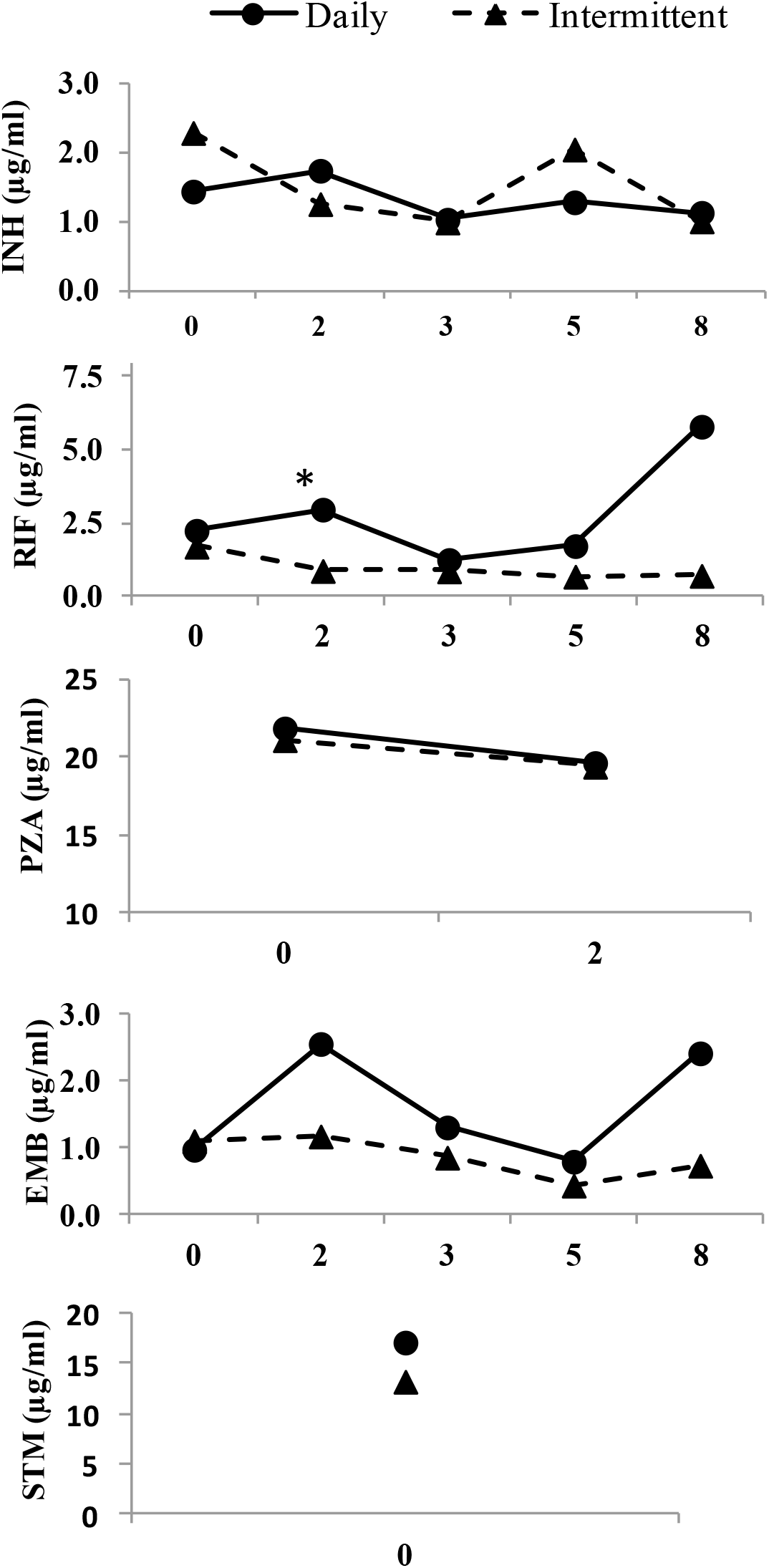
Daily vs intermittent regimen in PTB patients. Plasma levels of ATT drugs INH, RIF, PZA, EMB and STM at different time points of Daily vs Intermittent/Alternate Day regimen in PTB patients. X axis shows time points of drug administration i.e. day 0, 2 month, 3 month, 5 month and 8 month (0, 2, 3, 5 and 8).Y-axis shows Geometric Mean of plasma drug concentration (µg/ml) as measured by LC-MS in patient blood samples after 2hrs of drug ingestion. Solid line 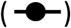 represents plasma levels of daily regimen subjects while intermittent regimen patient data is shown by a dashed line 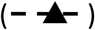.**\*** denotes statistical significance p≤ 0.05 Pyrazinamide is shown for upto 2 months and streptomycin C_2hr_ levels are shown for Day0

## Discussion

There is a paucity of studies discussing adverse outcomes in PTB re-treatment subjects. Here, we report a serial estimation of levels of first-line ATT drugs measured at five time-points during therapy and correlated to disease outcome exclusively in 134 PTB re-treatment patients.

We found a subject-wise variability in plasma concentrations of all four TB drugs which is in concordance with many other studies.(9, 15). Majority (68.5%) patients had lower drug concentrations than the reference values(15, 16). In the ‘Not Cured’ category, C_2hr_ levels of four ATT drugs remained much below reference target ranges for adults almost at all time points during the therapy, and were significantly lower than levels in ‘Cured’ patients. Patients with extremely low C_2hr_ ATT levels in plasma on day0 had outcomes such as acquired MDR-TB and treatment failure. Post-hoc analysis showed that patients with statistically significant low drug levels (65%) got acquired drug resistance within three months of enrolment in the study. All of these patients had lower levels in comparison to both cured and not cured patients (Figure 1b). No statistically significant difference in the drug plasma levels of patients on daily dose as compared to those on intermittent dosing was evident; however, the plasma levels were higher in daily dose patients (Figure4). A significant increase in microbiologically defined adverse outcomes in patients on intermittent therapy has been previously reported by Johnston et al. where pooled analysis and meta regression showed that intermittent regimen in ATT was inferior in all 3 outcomes viz. failure, relapse and acquired drug resistance (17). WHO has since 2008 recommended daily dosing (2) and country programs too has now implemented daily DOTS therapy (18)

Efficacy of first-line ATT has been shown to strongly associate with area under the curve (AUC). AUC derived from 9-time points sampling during the first 24 hrs post-drug administration were predictive of long-term clinical outcomes in PTB patients (12). Microbicidal effects of ATT are best described by AUC:MIC and Cmax:MIC ratios (19).Such studies require multiple sampling, are stressful to TB patients and not practical in resource-limited settings. We collected samples at trough concentrations (C_0_) and 2 hour concentrations(C_2hr_) post drug on day0, months 2, 3, 5 and 8 in order to identify the best indicator time point for single sampling as drug levels at C_2hr_ correlate well with Cmax and/or AUC (15, 16, 20, 21). However, the possibility that low C2_hr_ plasma concentrations could be a result of delayed absorption rather than a low peak concentration and could miss the actual peak concentration Cmax (21) cannot be ruled out.

Instead of using predetermined C_max_ concentrations, we first enrolled patients randomly and dichotomized them based on treatment outcomes (clinical with microbiological findings) at the end of the study. Loss to follow-up in a few patients, inability to provide sputum specimens, undetectable plasma drug levels etc. have caused a few gaps in our data. Patients were encouraged to comply and we sought to include all available data for analysis.

Peak drugs levels attained for INH, RIF and PZA reduced with time in our patients in both ‘cured’ and ‘not cured’ groups, possibly due to individual pharmacokinetic variations, poor drug absorption, pre-hepatic metabolism and hepatic enzymes induction, drug binding to plasma protein, or drug-drug interactions (22, 23). Rifampicin is reported to be highly bound to plasma proteins in TB patients as well as healthy subjects, leading to a loss in circulating levels, explaining decrease in bioavailability of Rifampicin from 93% to 68% after three weeks of treatment (24-26). It is tempting to hypothesize that drug-drug interaction is at its minimal on Day0, but may have stronger and complex interactions to compromise drug bioavailability resulting in lower C_2hr_ concentrations subsequently during therapy possibly due to induction of drug metabolism or transporter protein synthesis. Day0-C_2hr_ drug levels for cured patients were higher than subsequent time points except for EMB for which the levels were relatively constant.

We also report *therapeutic indicator levels*, the minimal drug plasma level values predicting a positive treatment outcome (Table 4) for all the four ATT drugs. We believe that the day0-C_2hr_ levels especially that of PZA might be useful as a cut-off reference for dosage adjustments for clinicians during therapy and help mitigate adverse events or treatment failure in DOTS Category II patients early on. Similar to our findings, a recent study where drug exposure and MIC were evaluated in PTB patients, PZA levels were shown to be important in predicting poor clinical response suggesting the use of PZA and RIF at levels above a threshold value to improve treatment outcomes. However, the inter-play between PZA and RIF may affect the final levels detected.(27)

Evaluation of certain other factors which may contribute to patient outcome was beyond the scope of the present study. Improvement in nutritional status and immune response may complement the pharmacotherapy. Low INH concentrations observed in the patients could also be due to the well recognised pharmacogenetic variants with acetylator status (28-30). Another limitation was that we could not delineate the cumulative effects of the drugs or drug– drug interference in the regimen. Our study had some missing data due to missing patient visits (follow-up) and few undetectable drug levels in some patients. These, however, will unlikely impact the overall results obtained.

Overall, we conclude that treatment outcomes in pulmonary tuberculosis patients on retreatment regimen are strongly linked to plasma levels of drugs, with low concentrations leading to poor treatment outcomes. Our findings emphasize the importance of therapeutic drug monitoring and dose adjustment to maintain plasma levels above a critical threshold.

## Methods and Materials

### Ethics

This study was done at the All India Institute of Medical Sciences (AIIMS), New Delhi, India from 2012-2014 and approved by the Institutional Ethics Committee, AIIMS (IEC/NP-251/2012&RP-22/2012)

### Patients

Patients diagnosed with drug susceptible PTB (and no other co-morbidities), aged between 15 to 65 yrs and advised the retreatment regimen under National TB Elimination Program (NTEP) (erstwhile Revised National TB Control Program (RNTCP) were included in this study (31). Informed written consent was obtained from each patient.

A detailed demographic data and clinical history was obtained (Table 1), and results of sputum acid-fast stains, cultures and all laboratory data were recorded (Table 2). A brief diagram of the study approach is shown in supplementary figure S1.

### Treatment Regimen

ATT was given as per the prevailing guidelines of the RNTCP at the time of the study (2012-2014) for previously treated TB cases (erstwhile Category II) i.e., INH + RIF+ EMB + PZA + injectable STM for 2 months, followed by INH + RIF+ EMB + PZA for 1 month, and INH + RIF+ EMB for 5 months as per the weight bands.(H,5mg/Kg, R,10 mg/Kg, Z,30 mg/Kg, E,15 mg/Kg, S,15 mg/Kg(in daily regimen) and H,10 mg/Kg, R,10 mg/Kg, Z,30 mg/Kg, E,15 mg/Kg, S,15 mg/Kg (intermittent regimen). RNTCP followed intermittent regimen recommended by WHO (2),Clinical follow-up of patients was done at 0 day, weeks 2, 4, 8, 12, 20, and 32 (at completion of 8 months). Therapy was not modified during the study.

### Sample collection

#### Sputum samples

For AFB smear, MGIT culture and Xpert MTB/RIF assays, 10 ml sputum sample was collected from study subjects at the time of recruitment (Day0) and at the end of 2, 3, 5 and 8 months.

#### Blood samples

Half ml blood sample was collected for the measurement of baseline drug concentration just before intake of daily dose of ATT and define the trough of drug level, on all days of sample collection. ATT medicines were given under observation of the field worker before breakfast as prescribed, following which the second blood sample (0.5 ml) was collected at 2 hours from the time of drug intake. Blood samples were collected in polypropylene BD vacuutainers containing EDTA and sodium ascorbate. Separated plasma was stored in plain endotoxin free vials at -80^°^C for further use.

#### Samples processing and AFB-staining

Samples were processed for decontamination by Modified Petroff’s method and stained for acid fast bacilli by Ziehl-Neelson’s staining as per standard protocol (32)

#### Culture for *Mycobacterium tuberculosis*

Concentrated sediment from decontaminated sample was inoculated in MGIT 960 medium (BD Biosciences) and cultured for *Mycobacterium tuberculosis* (MTB) using standard MGIT^™^ method as per manufacturer’s instructions (Beckton Dickinson, MD, USA)

### Xpert MTB/RIF Assay

Xpert MTB/RIF assay (Cepheid, Sunnyvale, CA, USA) was used for detection of MTB and resistance to Rifampicin, as per manufacturer’s instructions.

### LC-MS/MS and C_max_determination

Level of drugs in plasma samples were analysed by Liquid chromatography-tandem mass spectrometry (LC-MS/MS) as described previously (33). An Ultra High Performance Liquid Chromatographic system (UHPLC, Thermo Surveyor system, CITY, USA) coupled with a triple quadrupole mass spectrometer API-4000 QTrap fitted with Turbo Ion Spray Ionization (ESI) source (MDS SCIEX, Applied Biosystems, USA) was operated in positive ion mode for separation and detection of drug concentrations of the anti-tuberculosis drugs (H, R, E, Z and S). Standards were procured from Sigma-Aldrich Ltd, India and the validation of the analytical method was done as per United States Food and Drug Administration (US-FDA) guidelines for bioanalytical method validation (U.S. Food & Drug Administration, 2013).

Pharmacokinetic parameters of C_0hr_ were measured for baseline plasma drug concentration and C_2hrs_ for steady state C_max_ plasma concentration. The 2-hour plasma concentrations were compared with the reference C_2hr_ concentrations as described previously (15, 16)

### Evaluation of Treatment Response

Patients were defined as ‘cured’ if their sputum were AFB-negative within 5 months of treatment and continued to be negative until the end of treatment along with the physician’s clinical assessment of TB free status. ‘Not Cured’ status was defined as presence of a positive AFB smear beyond 5 months of a compliant tuberculosis treatment, presence of a drug-resistant strain during any point of treatment, or a physician’s evaluation of symptoms and clinical status after 5 months of treatment compared with the status at diagnosis. (31)

### Statistical Analysis

Data were recorded and analysed using MS-Excel-2007 and SPSS 22.0 for Windows® (SPSS Inc., Chicago, IL, USA).Chi-square tests were used to investigate any association of patient demography with clinical outcomes. Geometric mean (GM) plasma concentrations between the patients groups were compared using one way ANOVA and Mann Whitney U tests. Receiver operator characteristics (ROC) curves were used to study the discriminatory abilities of ATT plasma levels between the two categories and optimal cut off for therapeutic indicator levels (TIL). All tests were two sided, with P ≤ 0.05 indicating a statistically significant difference.

## Data Availability

All Raw data is available with the Principal investigator /corresponsding author of the research study.

## Acknowledgement

We are thankful to our patients for the success of this study. We also thank Mr. Ashish Datt Upadhayay, Statistical Assistant, Department of Biostatistics, AIIMS, New Delhi for the ROC curve analysis.

We thank DST-FIST for providing liquid chromatography coupled tandem mass spectroscopy facility for the analysis.

## Figure legends

**Figure S1:**
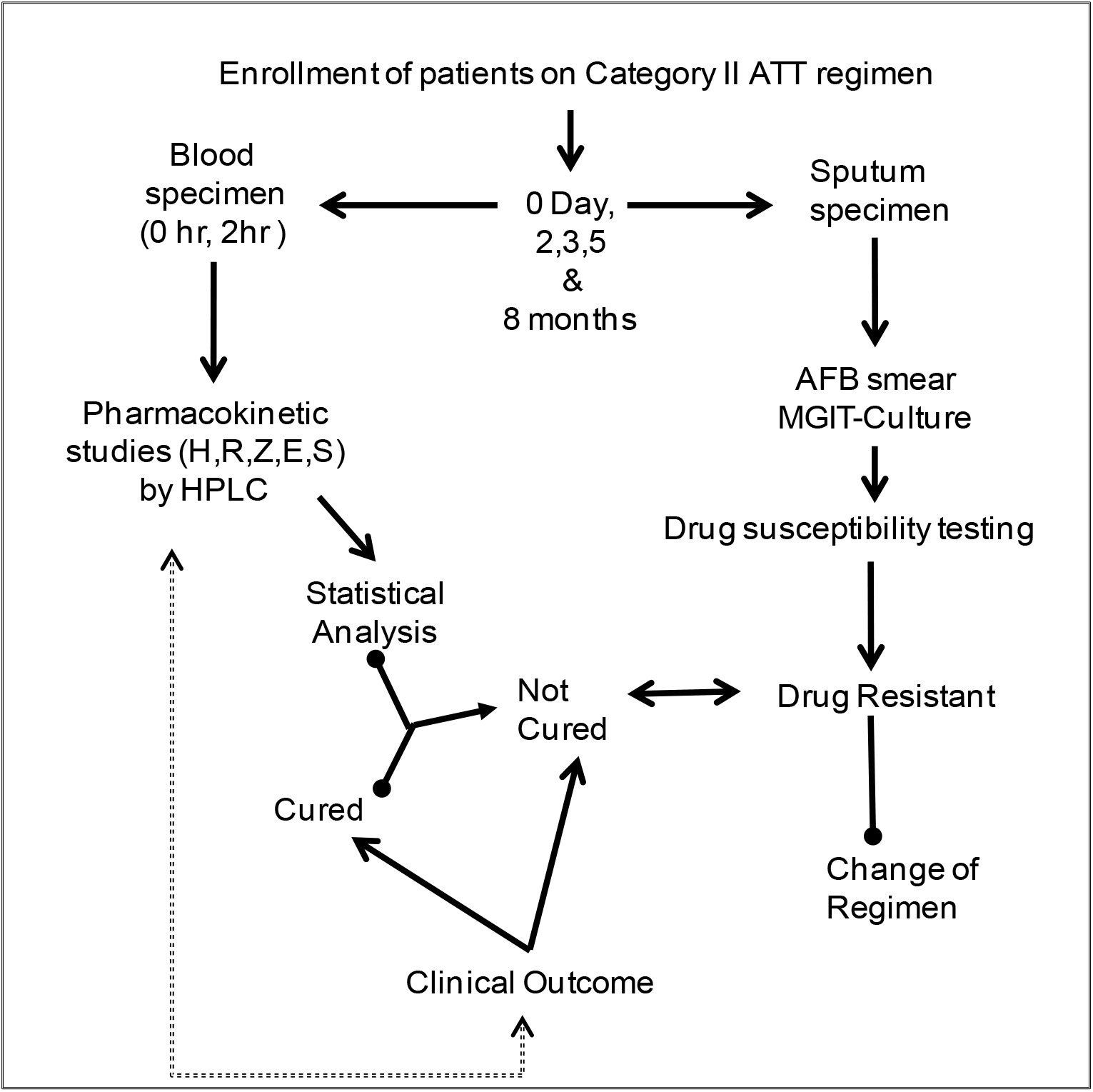
Study Approach: Figure illustrates a schematic design of the study from patient enrolment until statistical analysis

## References

1. WHO. 2013; updated Dec 2014 and Jan 2020. Definitions and reporting framework for tuberculosis—2013 revision. World Health Organisation Switzerland, Geneva.

2. WHO. 2010 (updated 2017). Treatment of Tuberculosis Guidelines,4th edition Organisation WH, Geneva, Switzerland.

3. Jones-López EC, Ayakaka I, Levin J, Reilly N, Mumbowa F, Dryden-Peterson S, Nyakoojo G, Fennelly K, Temple B, Nakubulwa S, Joloba ML, Okwera A, Eisenach KD, McNerney R, Elliott AM, Ellner JJ, Smith PG, Mugerwa RD. 2011. Effectiveness of the standard WHO recommended retreatment regimen (category II) for tuberculosis in Kampala, Uganda: a prospective cohort study. PLoS Med 8:15.

4. Cohen DB, Davies G, Malwafu W, Mangochi H, Joekes E, Greenwood S, Corbett L, Squire SB. 2019. Poor outcomes in recurrent tuberculosis: More than just drug resistance? PLoS One 14.

5. Sisodia RS, Wares DF, Sahu S, Chauhan LS, Zignol M. 2006. Source of retreatment cases under the revised national TB control programme in Rajasthan, India, 2003. Int J Tuberc Lung Dis 10:1373–9.

6. Kimerling ME, Phillips P, Patterson P, Hall M, Robinson CA, Dunlap NE. 1998. Low serum antimycobacterial drug levels in non-HIV-infected tuberculosis patients. Chest 113:1178–83.

7. Barakat MT, Scott J, Hughes JM, Walport M, Calam J, Friedland JS, Ind PW, McKenna C. 1996. Grand rounds--Hammersmith Hospital. Persistent fever in pulmonary tuberculosis. Bmj 313:1543–5.

8. Heysell SK, Moore JL, Keller SJ, Houpt ER. 2010. Therapeutic drug monitoring for slow response to tuberculosis treatment in a state control program, Virginia, USA. Emerg Infect Dis 16:1546–53.

9. Chideya S, Winston CA, Peloquin CA, Bradford WZ, Hopewell PC, Wells CD, Reingold AL, Kenyon TA, Moeti TL, Tappero JW. 2009. Isoniazid, rifampin, ethambutol, and pyrazinamide pharmacokinetics and treatment outcomes among a predominantly HIV-infected cohort of adults with tuberculosis from Botswana. Clin Infect Dis 48:1685–94.

10. Park JS, Lee JY, Lee YJ, Kim SJ, Cho YJ, Yoon HI, Lee CT, Song J, Lee JH. 2015. Serum Levels of Antituberculosis Drugs and Their Effect on Tuberculosis Treatment Outcome. Antimicrob Agents Chemother 60:92–8.

11. Srivastava S, Pasipanodya JG, Meek C, Leff R, Gumbo T. 2011. Multidrug-resistant tuberculosis not due to noncompliance but to between-patient pharmacokinetic variability. J Infect Dis 204:1951–9.

12. Pasipanodya JG, McIlleron H, Burger A, Wash PA, Smith P, Gumbo T. 2013. Serum drug concentrations predictive of pulmonary tuberculosis outcomes. J Infect Dis 208:1464–73.

13. Zuur MA, Bolhuis MS, Anthony R, den Hertog A, van der Laan T, Wilffert B, de Lange W, van Soolingen D, Alffenaar JW. 2016. Current status and opportunities for therapeutic drug monitoring in the treatment of tuberculosis. Expert Opin Drug Metab Toxicol 12:509–21.

14. Motta I, Calcagno A, Baietto L, Bigliano P, Costa C, Baruffi K, Fatiguso G, D’Avolio A, Di Perri G, Bonora S. Pharmacokinetics of first-line antitubercular drugs in plasma and PBMCs. Br J Clin Pharmacol. 2017 May;83(5):1146–1148. doi: 10.1111/bcp.13196. Epub 2017 Jan 18.

15. Peloquin CA. 2002. Therapeutic drug monitoring in the treatment of tuberculosis. Drugs 62:2169–83.

16. Burhan E, Ruesen C, Ruslami R, Ginanjar A, Mangunnegoro H, Ascobat P, Donders R, van Crevel R, Aarnoutse R. 2013. Isoniazid, rifampin, and pyrazinamide plasma concentrations in relation to treatment response in Indonesian pulmonary tuberculosis patients. Antimicrob Agents Chemother 57:3614–9.

17. Johnston JC, Campbell JR, Menzies D. 2017. Effect of Intermittency on Treatment Outcomes in Pulmonary Tuberculosis: An Updated Systematic Review and Metaanalysis. Clin Infect Dis 64:1211–1220.

18. NTEP (National Tuberculosis Elimination Programme fRNTCP, RNTCP). 2017 March. National Strategic Plan for Tuberculosis: 2017-25 Elimination by 2025. Welfare MoHwF, Ministry of Health with Family Welfare, Nirman Bhawan, New Delhi. https://tbcindia.gov.in/WriteReadData/National%20Strategic%20Plan%202017-25.pdf.

19. Hall RG, Leff RD, Gumbo T. 2009. Treatment of active pulmonary tuberculosis in adults: current standards and recent advances. Insights from the Society of Infectious Diseases Pharmacists. Pharmacotherapy 29:1468–81.

20. Saktiawati AMI, Harkema M, Setyawan A, Subronto YW, Sumardi Stienstra Y, Aarnoutse RE, Magis-Escurra C, Kosterink JGW, van der Werf TS, Alffenaar JC, Sturkenboom MGG. 2019. Optimal Sampling Strategies for Therapeutic Drug Monitoring of First-Line Tuberculosis Drugs in Patients with Tuberculosis. Clin Pharmacokinet 58:1445–1454.

21. Peloquin CA. 1997. Using therapeutic drug monitoring to dose the antimycobacterial drugs. Clin Chest Med 18:79–87.

22. Loos U, Musch E, Jensen JC, Schwabe HK, Eichelbaum M. 1987. Influence of the enzyme induction by rifampicin on its presystemic metabolism. Pharmacol Ther 33:201–4.

23. Loos U, Musch E, Jensen JC, Mikus G, Schwabe HK, Eichelbaum M. 1985. Pharmacokinetics of oral and intravenous rifampicin during chronic administration. Klin Wochenschr 63:1205–11.

24. Peloquin CA. 2001. Pharmacological issues in the treatment of tuberculosis. Ann N Y Acad Sci 953:157–64.

25. Nahid P, Dorman SE, Alipanah N, Barry PM, Brozek JL, Cattamanchi A, Chaisson LH, Chaisson RE, Daley CL, Grzemska M, Higashi JM, Ho CS, Hopewell PC, Keshavjee SA, Lienhardt C, Menzies R, Merrifield C, Narita M, O’Brien R, Peloquin CA, Raftery A, Saukkonen J, Schaaf HS, Sotgiu G, Starke JR, Migliori GB, Vernon A. 2016. Official American Thoracic Society/Centers for Disease Control and Prevention/Infectious Diseases Society of America Clinical Practice Guidelines: Treatment of Drug-Susceptible Tuberculosis. Clin Infect Dis 63:e147–e195.

26. Boman G, Ringberger VA. 1974. Binding of rifampicin by human plasma proteins. Eur J Clin Pharmacol 7:369–73.

27. Zheng X, Bao Z, Forsman LD, Hu Y, Ren W, Gao Y, Li X, Hoffner S, Bruchfeld J, Alffenaar JW. 2020. Drug exposure and minimum inhibitory concentration predict pulmonary tuberculosis treatment response. Clin Infect Dis 18.

28. Meyer UA, Zanger UM. 1997. Molecular mechanisms of genetic polymorphisms of drug metabolism. Annu Rev Pharmacol Toxicol 37:269–96.

29. Tostmann A, Mtabho CM, Semvua HH, van den Boogaard J, Kibiki GS, Boeree MJ, Aarnoutse RE. 2013. Pharmacokinetics of first-line tuberculosis drugs in Tanzanian patients. Antimicrob Agents Chemother 57:3208–13.

30. Aarnoutse R. 2011. Chapter 18: Pharmacogenetics of Antituberculosis Drugs, vol vol 40. Basel, Karger.

31. NTEP (National Tuberculosis Elimination Programme fRNTCP, RNTCP). 2016. Technical and Operational Guidelines for TB Control in India Directorate General of Health Services MoHaFW, Nirman Bhawan, Central TB Division, New Delhi - 110001.

32. RNCTP. April 2009. Training Manual for Mycobacterium tuberculosis Culture & Drug susceptibility testing. Directorate General of Health Services MoHaFW, Nirman Bhawan, Ministry of Health and Family Welfare, Nirman Bhawan,New Delhi 110011,

33. Mukherjee A, Velpandian T, Singla M, Kanhiya K, Kabra SK, Lodha R. 2016. Pharmacokinetics of isoniazid, rifampicin, pyrazinamide and ethambutol in HIV-infected Indian children. Int J Tuberc Lung Dis 20:666–72.

